# Missing data matters in participatory syndromic surveillance systems: comparative evaluation of missing data methods when estimating disease burden

**DOI:** 10.1101/2021.05.11.21256420

**Authors:** Kristin Baltrusaitis, Craig Dalton, Sandra Carlson, Laura F. White

## Abstract

**Introduction:** Traditional surveillance methods have been enhanced by the emergence of online participatory syndromic surveillance systems that collect health-related digital data. These systems have many applications including tracking weekly prevalence of Influenza-Like Illness (ILI), predicting probable infection of Coronavirus 2019 (COVID-19), and determining risk factors of ILI and COVID-19. However, not every volunteer consistently completes surveys. In this study, we assess how different missing data methods affect estimates of ILI burden using data from FluTracking, a participatory surveillance system in Australia.

**Methods:** We estimate the incidence rate, the incidence proportion, and weekly prevalence using five missing data methods: available case, complete case, assume missing is non-ILI, multiple imputation (MI), and delta (*δ*) MI, which is a flexible and transparent method to impute missing data under Missing Not at Random (MNAR) assumptions. We evaluate these methods using simulated and FluTracking data.

**Results:** Our simulations show that the optimal missing data method depends on the measure of ILI burden and the underlying missingness model. Of note, the *δ*-MI method provides estimates of ILI burden that are similar to the true parameter under MNAR models. When we apply these methods to FluTracking, we find that the *δ*-MI method accurately predicted complete, end of season weekly prevalence estimates from real-time data.

**Conclusion:** Missing data is an important problem in participatory surveillance systems. Here, we show that accounting for missingness using statistical approaches leads to different inferences from the data.

## INTRODUCTION

Over the past two decades, traditional surveillance methods have been enhanced by the emergence of online participatory syndromic surveillance systems that collect health-related digital data in near-real time.[1] Through these systems, participants volunteer to regularly report syndromic, health information via online or mobile communication technologies. These systems complement traditional healthcare-based surveillance systems by reducing the time delay associated with visiting a healthcare provider and capturing individuals who do not seek medical care.[2] The first of these systems, de Grote Griepmeting, or the Great Influenza Survey, started in 2003 in the Netherlands and Belgium,[3] and multiple systems throughout Europe,[4] Australia (AU),[5] the United States (US),[6] Mexico, and Brazil have followed. More recently, participatory syndromic surveillance systems have been deployed to track symptoms of Coronavirus 2019 (COVID-19) in the community.[7]

Participatory syndromic surveillance systems are being used to forecast weekly prevalence of Influenza-Like Illness (ILI),[9–13] produce age-specific attack rates of ILI,[14–16] predict probable infection of COVID-19,[17] determine risk factors of ILI and COVID-19,[11,17] estimate influenza vaccine effectiveness,[11,18,19] and assess healthcare-seeking behavior.[20-22] methods to FluTracking, we find that the *δ*-MI method accurately predicted complete, end of These systems actively engage the public in reporting and providing timely information about disease trends within the community, while providing a mechanism for community members to become “citizen-scientists”.[2,8] However, because not every participant reports regularly the population at risk can be temporally inconsistent and include systematic biases.[23] For example, in post-influenza season surveys, over 30% of users from the U.S. system, Flu Near You (FNY), reported they were at least slightly more likely to report if they have had symptoms. Furthermore, during the influenza season, FNY users were significantly less likely to consistently submit reports if they reported symptoms during their first report following registration.[24] Of note, the reporting habits of users vary by system. For example, approximately 22% of FNY users submitted at least half of the symptom reports during the 2013-2014 influenza season,[6] whereas the AU system, FluTracking, reported that during the 2017 influenza season, of the participants who completed a survey during the first four survey weeks, 69% completed all available surveys, and 82% completed over 90% of available surveys.[5]

The most common approach for addressing the inconsistencies in the user reporting is to select a cohort of “active users”, where the definition of “active user” varies by system and study, and assume that all missing reports were asymptomatic.[11,14–16] However, no study has assessed how this deterministic assumption affects estimates or systematically compared approaches through simulations.

Here, we draw upon statistical methods for missing data and assess how different missing data methods, such as complete case and imputation-based methods, affect estimates of ILI burden using both simulated data and data from FluTracking.

## METHODS

### FluTracking

FluTracking is an online health surveillance system of influenza in AU and, as of 2018, New Zealand and Hong Kong in 2021. Launched in 2006, the FluTracking system has grown to include over 150,000 participants during the peak weeks of COVID-19 surveillance across Australia and New Zealand in 2020.[25] At registration, FluTracking users provide basic demographic information, including month and year of birth, sex, postcode of residence, Indigenous status, highest level of education, and whether or not they work directly with patients in a healthcare setting. After registration, users complete weekly surveys for themselves and other household members about ILI symptoms including fever, cough, and/or sore throat. Users who report any symptoms are asked follow-up questions about absenteeism from work or normal duties, visits to health care providers, and results of laboratory tests. All users are also asked about influenza vaccination. Symptom surveys are sent every Monday, however, unlike other participatory surveillance systems, FluTracking participants have the option to complete missed surveys up to five weeks previous, called “retrospective reports”.

For this study, ILI is defined as report of both fever and cough, with or without sore throat. Descriptive statistics for participant characteristics, including age, sex, household status, and influenza vaccination status, are displayed as median (25^th^ percentile, 75^th^ percentile) for continuous variables and n (%) for categorical variables for of all participants who submitted at least one symptom report during the 2016, 2017, and 2018 influenza seasons. Although FluTracking collects data from the beginning of May through mid-October, we use reports submitted during the southern hemisphere’s influenza surveillance season, defined as Morbidity and Mortality weeks 25 through 41, or approximately late-June through mid-October.

### Measures of Influenza-Like Illness Burden

As recommended by the World Health Organization (WHO), we use the incidence rate (IR) as one measure of ILI burden.[26] The IR is equal to the number of incident ILI reports, defined as a report of ILI in which ILI was not reported the previous week, divided by the total person-time reported by participants:

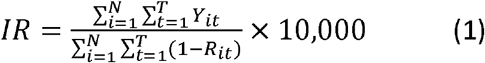

where

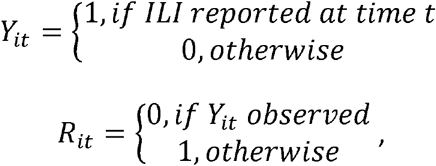

and t indexes the week numbers and ranges from 25 to 41.

The rate is expressed per 10,000 person weeks. Because person-time at risk is unavailable for most routine influenza surveillance data, we also present the incidence proportion (IP) for comparability across systems. The IP is equal to the number of participants who reported ILI at least once during the influenza season divided by the total number of participants:

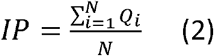

where

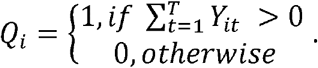

The 95% Confidence Intervals (CI) for these estimates are given by

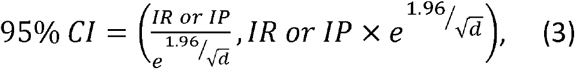

where *d* is the number of cases.[27,28] We calculate these measures of ILI burden for the overall population and by age group (<5, 5-17, 18-49, 50+ years). Finally, we present the weekly prevalence (WP) of ILI at each week, which is calculated by dividing the number of ILI reports by the total number of reports observed,

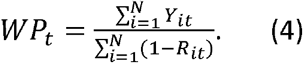

Because WP is estimated in near-real time, all retrospective reports were assumed missing when calculating estimates. We assess and compare these measures of ILI burden across three influenza seasons (2016, 2017, and 2018).

### Missing Data Methods

We assess five methods that account for missing data:

1. Available case
2. Complete case
3. Assume all missing reports are non-ILI
4. Multiple imputation (MI)
5. MI with delta (*δ*) adjustment

The first method uses all available cases (i.e., all reports submitted or select all *Y*_*it*_ for *R*_*it*_ =0), whereas the second method includes only complete cases (i.e., reports from individuals who submitted all reports or select all individuals, *i*, where *R*_*it*_ =0), for all *t*). These methods assume that data is Misssing Completely at Random (MCAR). In other words, the causes of the missing data are unrelated to the data. The third method assumes that all missing reports are non-ILI reports (i.e., *P*(*Y*_*it*_ =1 |*R*_*it*_ =1)=0), similar to past studies. The next two methods use MI methods to produce 10 point estimates, which are aggregated using Rubin’s rules with a log transformation to account for non-normality.[29] For the first MI method (method 4), we fit a model assuming Missing at Random (MAR),

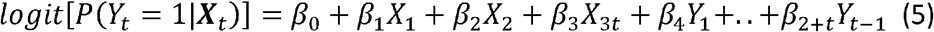

where

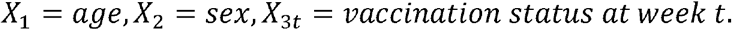

Under MAR, the probability that a value is missing depends only on the observed data and not the value itself. Because this assumption may not be valid for this data, we also perform MI using an *δ* adjustment, which is a flexible and transparent method to impute missing data under Missing Not at Random (MNAR) assumptions.[30] For MNAR, the probability that a value is missing depends on the unobserved data. The *δ*-MI method (method 5) uses (5), however, prior to imputing the missing data, a fixed quantity, *δ*, is added to the linear predictor of the regression model,

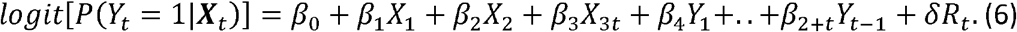

Here, *δ* represents the difference in log-odds of ILI between participants who did not report compared with participants who did report.

#### Estimation of delta

Because the log-odds of ILI for participants who did not report are unknown, we use the retrospective reports to estimate this value and create annual 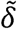 estimates. In other words,

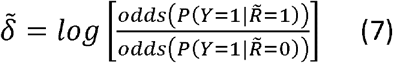

where,

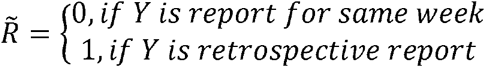

Negative values of 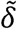 indicate that participants were less likely to report ILI for retrospective reports compared with reports submitted during the same week. Data are analyzed using R, version 3.3.2, and both MI methods are fit using the Multivariate Imputation by Chained Equations (MICE) package.[31,32]

### Simulations

We evaluate the missing data methods assuming three missingness scenarios: MCAR, MAR, and MNAR, using simulated data. The data is simulated using a three-step process. First, 1000 FluTracking populations (n=30,000 each) are simulated using the characteristics (age group, sex, and vaccination status) of the 2016 influenza season participant population. Simulated participants are assigned an age group, sex, vaccination status, and 17 weeks of symptom reports, Y_*it*_. These weekly symptom reports are simulated using a multinomial distribution, where n, which is Poisson distributed with an age-group specific mean, represents the total number of ILI reports for the participant and ***q*** = {*q*_1_, …,*q*_17_}, is the vector of weekly percentage of the 2016 sentinel general practitioner consultations that were ILI as reported by AU’s Department of Health.[33] We simulate 17 missingness indicators, *R*_*it*_, to reflect distribution of FluTracking participant reports (Supplemental Figure 1), using three missingness scenarios:

1. MCAR

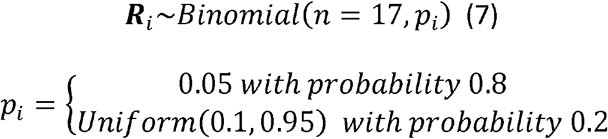
2. MAR

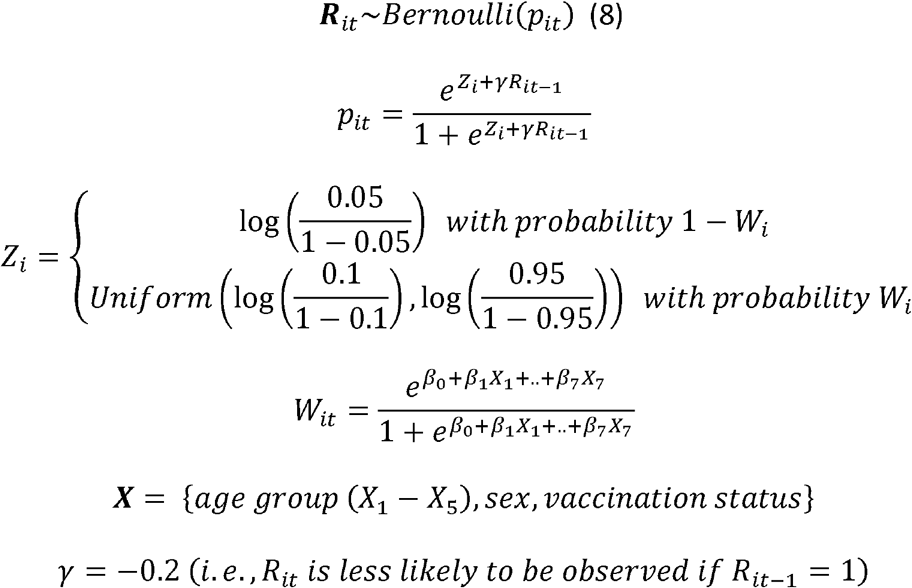
3. MNAR

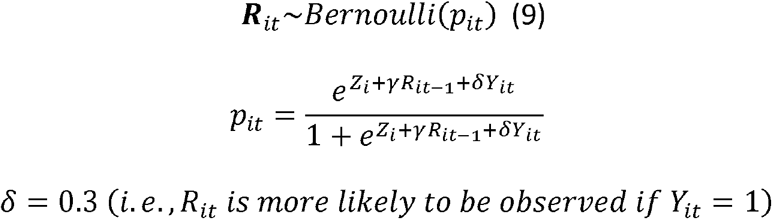

The values of *β*_0_ through *β*_7_ are estimated for FluTracking data using multivariate logistic regression, similar to [23]. We define *δ* equal to 0.3. However, we also present a sensitivity analysis that assesses how varying the MNAR assumption affects IR estimates (*δ* equal to 0.3 0.8, 1.3, 3.0, and 5.0). Finally, each of the missing data methods described in the previous section is applied to produce overall and age-specific IR and IP estimates, and the overall WP estimates for each simulated dataset. The estimates from each missing data method are compared to the original simulation parameters qualitatively using violin plots and quantitatively Normalized Root Mean Square Errors (NRMSE) normalized by the original parameter,

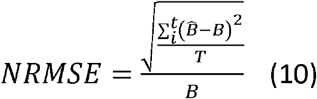

for T = 1000. In this equation, B represents the measure of ILI burden (i.e., IR, IP, or WP).

## RESULTS

### Simulations

#### Incidence Rate

As shown in Figure 1, under each missingness scenario, assuming that all missing reports are non-ILI underestimates the IR and results in the largest NRMSE (Table 1). Under MCAR and MAR scenarios, available case and MI methods have smaller NRMSEs compared with the complete case and *δ*-MI methods. However, under MNAR, IR estimates using the *δ*-MI method have smaller NRMSEs compared with all other methods.

**Table 1.**
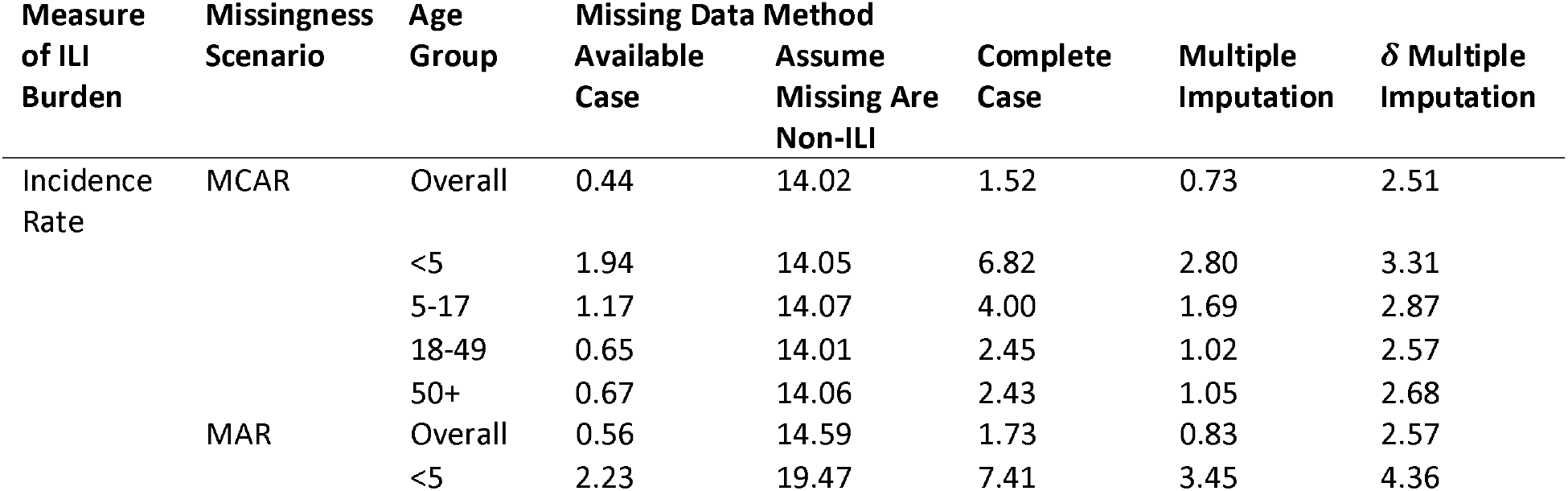
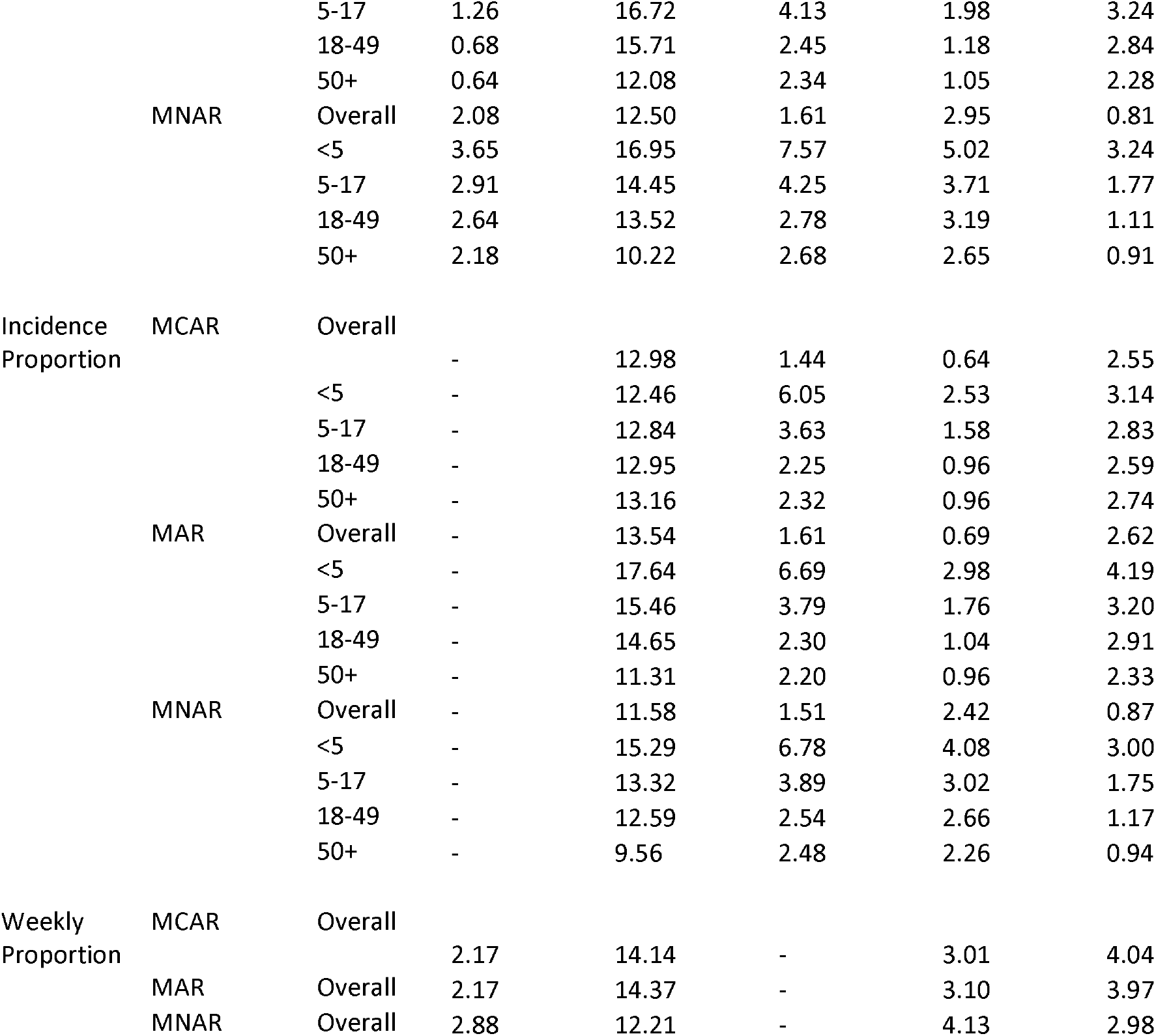
Normalized Root Mean Square Errors, expressed as percentage, by age group for Incidence Rates (IR) and Incidence Proportions (IP), and overall for Weekly Prevalence (WP) under Missing Completely at Random (MCAR), Missing At Random (MAR), and Missing Not at Random (MNAR) scenarios.

#### Incidence Proportion

Under all missingness scenarios, the complete case and imputation methods outperform the available case method (Figure 2, Table 1). For MCAR and MAR scenarios, the MI method has the smallest NRMSEs, the *δ*-MI method underestimates the IP, and the complete case method has the largest variation in estimates. Similar to IR, the *δ*-MI method is the best approach when data are MNAR. Estimates for the available case method are not shown because this method is the same as assuming that all missing reports are non-ILI when estimating the IP.

**Figure 1.**
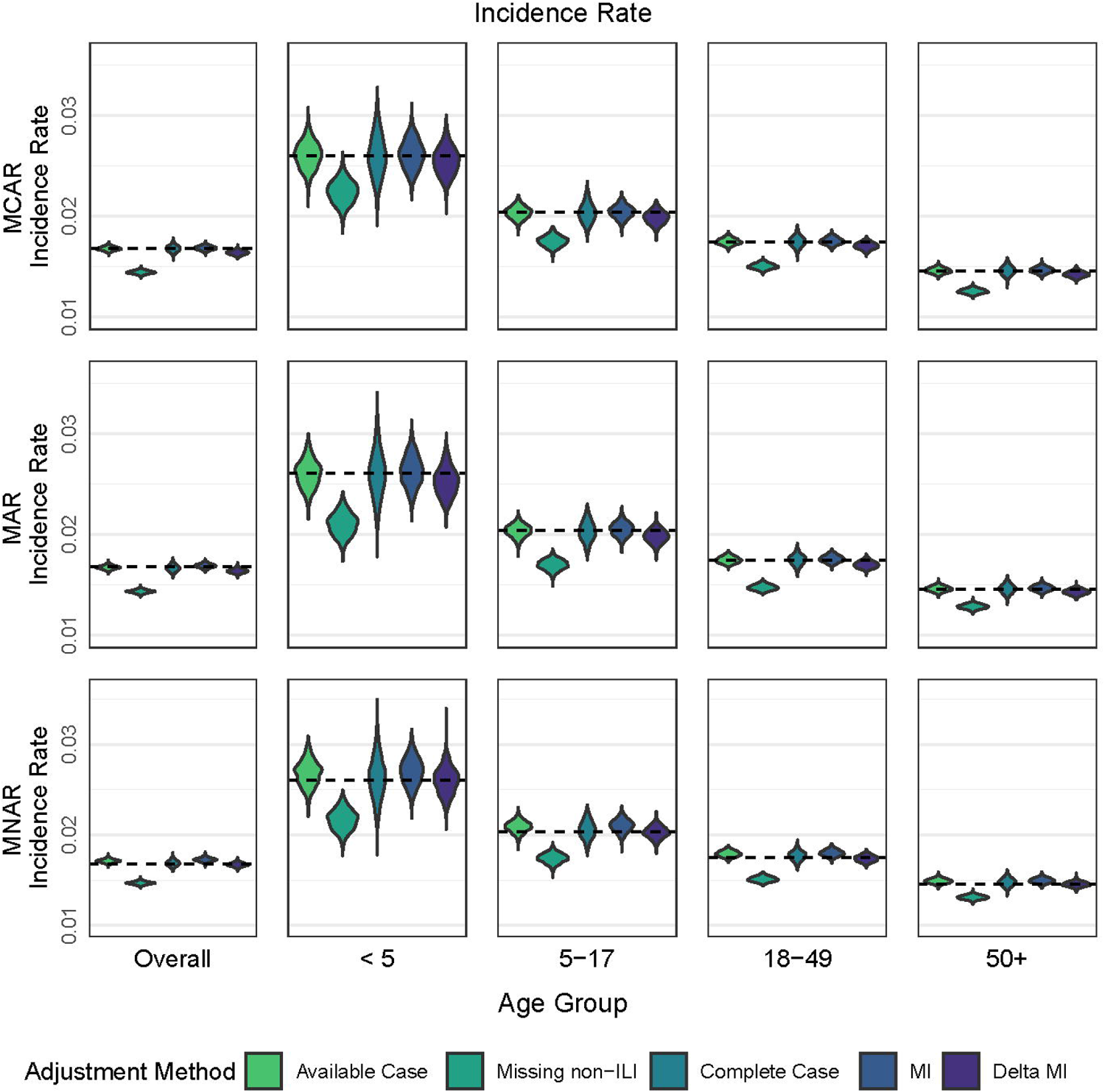
Distributions of overall and age-group specific Incidence Rates simulated under three missingness scenarios: Missing Completely at Random (MCAR), Missing at Random (MAR), and Missing Not at Random (MNAR). Dotted line represents the original simulated parameter.

**Figure 2.**
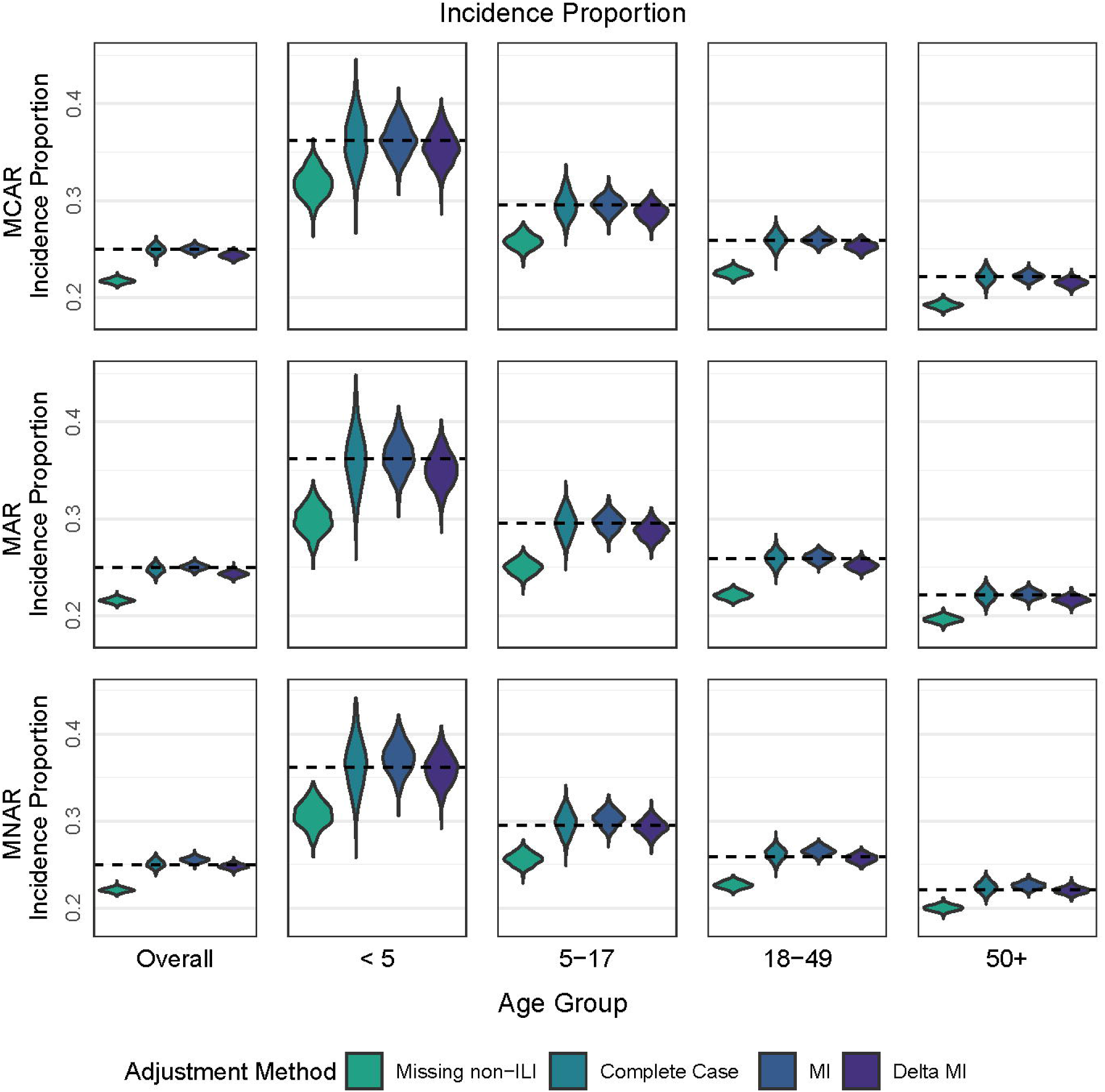
Distributions of overall and age-group specific Incidence Proportions simulated under three missingness scenarios: Missing Completely at Random (MCAR), Missing at Random (MAR), and Missing Not at Random (MNAR). Dotted line represents the original simulated parameter.

#### Weekly Prevalence

When estimating the WP, the available case method outperforms the other methods under each missingness scenario (Figure 3, Table 1). The method that assumes all missing reports are non-ILI underestimates the WP under each missingness scenario, and the *δ*-MI method outperforms MI when data are MNAR. Because the complete case population is unknown until the end of the season, we did not assess this missing data method.

**Figure 3.**
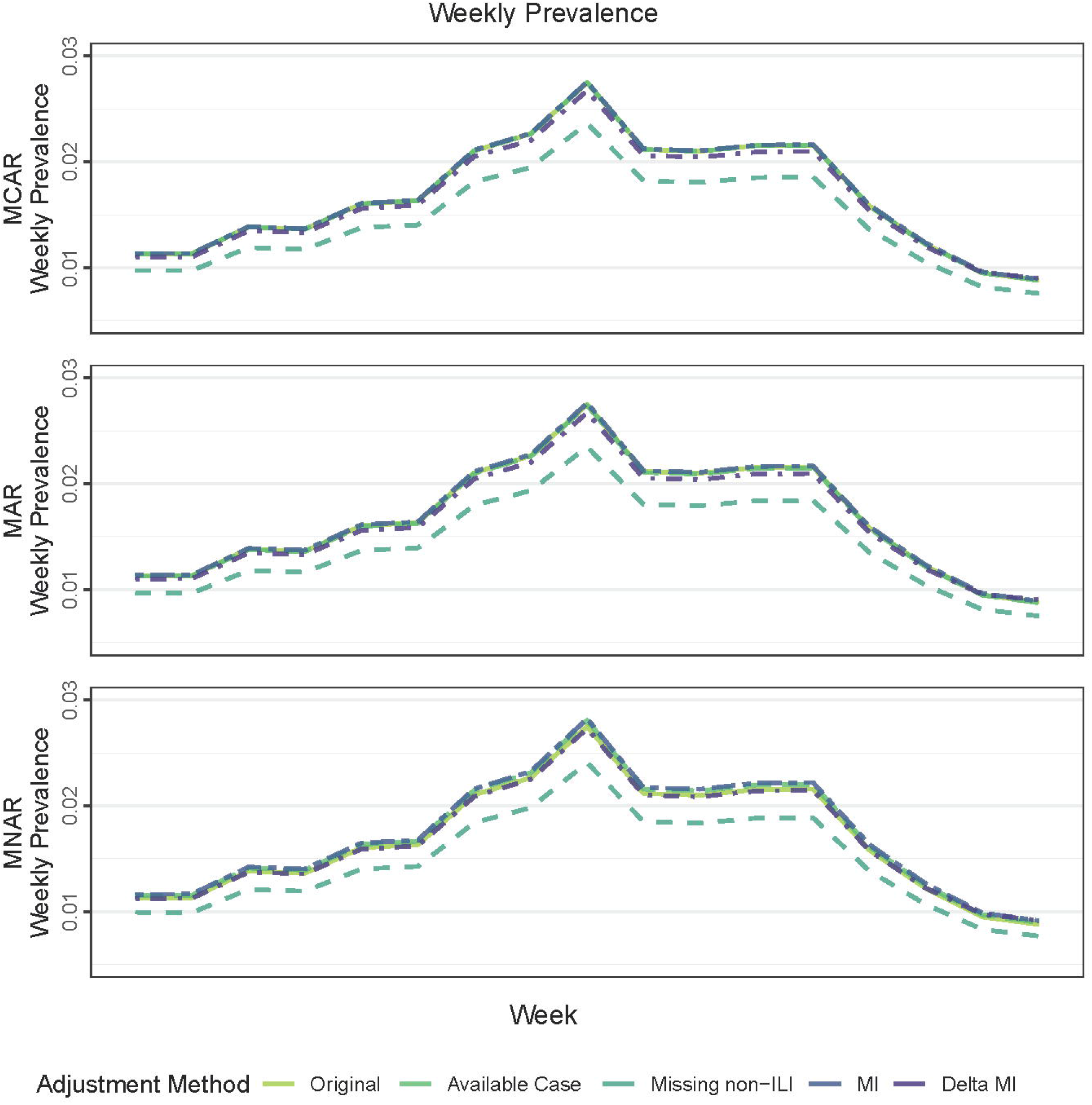
Time series of Weekly Prevalence simulated under three missingness scenarios: Missing Completely at Random (MCAR), Missing at Random (MAR), and Missing Not at Random (MNAR).

#### Sensitivity Analysis

Sensitivity Analysis results for calculation of overall and age group specific IRs under five MNAR scenarios are shown in Supplemental Figure 2. As the value of *δ* increases (i.e., ILI reports become less likely to be missing), the method that assumes missing reports are non-ILI becomes a better missing data method.

### FluTracking

During the 2016, 2017, and 2018 influenza seasons, 29,671, 32,778, and 43,389 unique participants submitted at least one symptom report between week 25 and week 41, respectively. Across all influenza seasons, approximately 60% identified as female, and the median age of participants ranged from 47 to 49 years. The largest age group was 50+ years, followed by 18 to 49, 5 to 17, and finally <5. More than half were primary users who submitted reports on their own behalf, and 59%, 61%, and 67% of participants reported that they received the influenza vaccination during the 2016, 2017, and 2018 influenza seasons, respectively (Table 2).

**Table 2.**
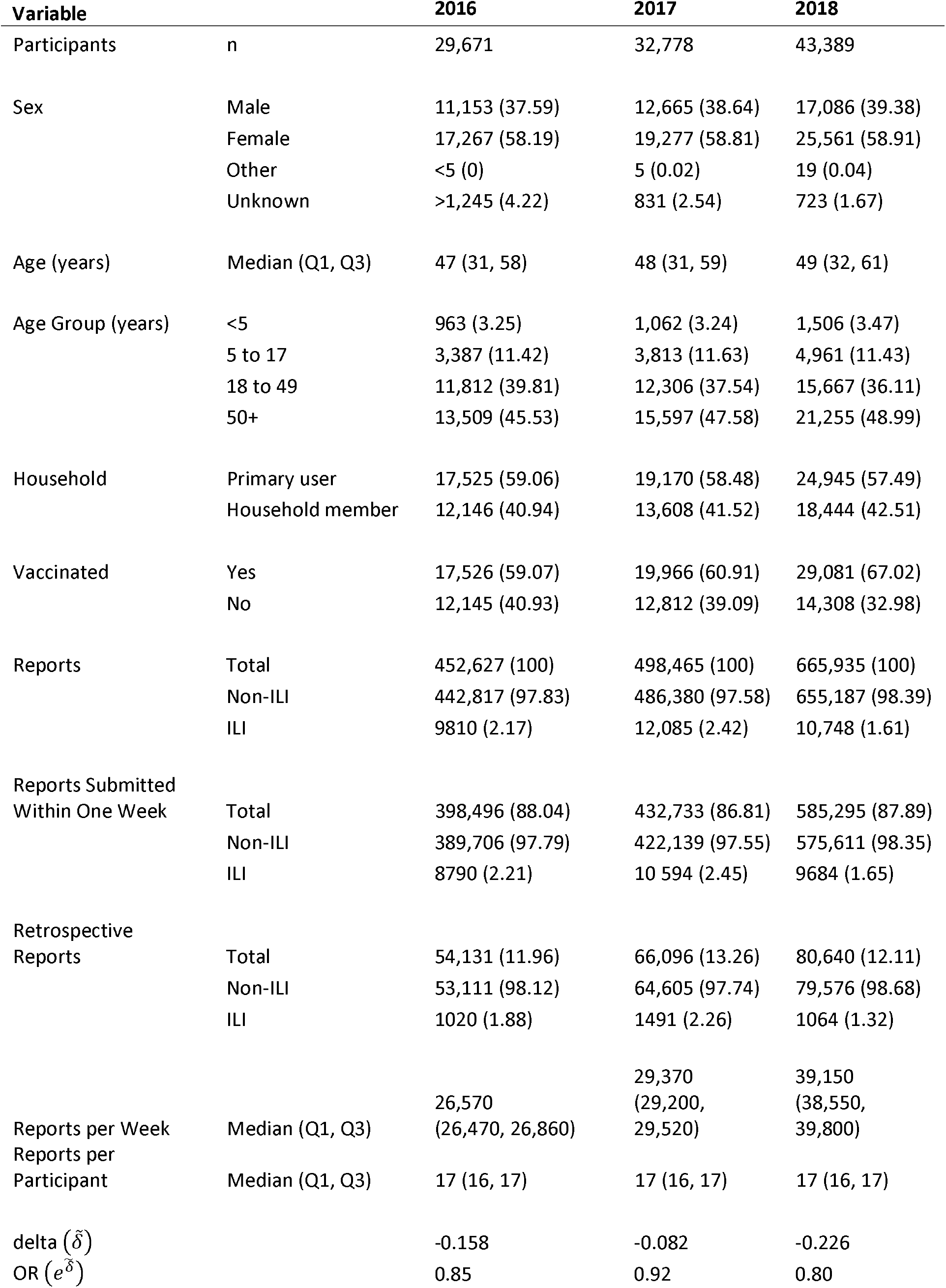
Descriptive statistics of the FluTracking cohort and participant reporting habits during the 2016, 2017, and 2018 influenza seasons. Continuous variables are displayed as median (25^th^ percentile, 75^th^ percentile) and categorical variables are displayed as n (%).

The descriptive statistics of the reporting habits of participants during the 2016, 2017, and 2018 influenza seasons are also shown in Table 2. The total number of symptom reports submitted during the influenza season increased from approximately 450,000 in 2016, to almost 500,000 in 2017, and finally to over 650,000 in 2018. The median number of weekly reports also increased from approximately 26,000 in 2016 to 39,000 in 2018. During each influenza season, the median number of reports per participant was 17 (16, 17), indicating that more than half of participants submitted a symptom report each week (Supplemental Figure 1). While most reports were submitted within one week of the symptom report date, a larger proportion of these reports were ILI compared to the retrospective reports, resulting in a negative 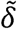. Although the exact value of 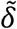 varies by season, the corresponding ORs of reporting ILI for a retrospective report compared with reporting ILI for a report submitted the same week are all less than 1. The OR was 0.85 in 2016, 0.92 in 2017, and 0.80 in 2018.

As shown in Supplemental Figure 1, most FluTracking participants register before week 25, however, the percentage of participants lost to follow-up increases as the season progresses, as shown by the bright green bars. The percentage of retrospective reports (dark green) is fairly consistent through the season, but the proportion of missing reports (light green) appears to increase until mid-season, at which point it slowly decreases as more participants are lost to follow-up (bright green). These patterns are consistent across all influenza seasons.

#### Incidence Rate

Supplemental Table 1 and Supplemental Figure 3 display overall and age-group specific IRs and 95% CIs, expressed as number of ILI reports per 10,000 person weeks, by influenza season. Although the 2017 influenza season had higher IRs compared with the 2016 and 2018 seasons, the general patterns in estimates are consistent across all seasons and age groups. The method that assumes that all missing reports are non-ILI has the lowest IR estimates, whereas available case and MI methods have the highest IR estimates. As expected, IR estimates from the *δ*-MI method are slightly less than estimates from the MI method without the *δ* adjustment, reflecting that missing reports are less likely to be ILI. In most age groups, IR estimates from the complete case method are similar or slightly greater than estimates from the method that assumes all missing reports are non-ILI.

#### Incidence Proportion

The overall and age-group specific IPs and 95% CIs for each influenza season are shown in Supplemental Table 2 and Supplemental Figure 4. Similar to IR estimates, IPs estimates from the method that assumes all missing reports are non-ILI and complete case method are less than IP estimates from the MI and *δ*-MI methods. However, the differences in IP estimates appear to be less pronounced compared with the differences in IR estimates.

#### Weekly Prevalence

Near-real time WP estimates are shown in Supplemental Figure 5. We also present the complete data with the retrospective reports for comparison. The method that assumes all missing reports are non-ILI results in WP estimates lower than the other methods. WP estimates from the MI method are slightly larger than WP estimates from the available case method and the *δ*-MI method. The estimates from these two methods are similar to the complete data.

## DISCUSSION

As we have shown, participatory surveillance systems, due to their design, suffer from substantial missing data that is likely informative. We have described five ways to handle missing data, drawing upon traditional statistical practice, and shown how these perform under different missingness scenarios. Our simulations show that the optimal missing data method depends on the measure of ILI burden and the underlying missingness model. Of note, the *δ*-MI method provides estimates of ILI burden that are similar to the true parameter under MNAR scenarios. When we apply these methods to a participatory surveillance system in Australia, we find that the *δ*-MI method accurately predicted end of season WP estimates from real-time data.

In 2020, Liu et al. estimated WP of ILI in AU using a Bayesian approach that adjusts FluTracking estimates by the probability of an individual reporting in each week given their past reporting.[34] Similar to our approach, they show that estimates of ILI burden are affected by models that correct for user behavior and that users are more likely to report when ill. However, their weekly behavior-adjusted estimates of prevalence were less than the WP estimates that assume all missing reports were non-ILI during the 2017 influenza season. This difference may indicate that their model assumes that the weekly percentage of ILI in the FluTracking population is greater than the weekly percentage of ILI in the general AU population. Their approach also differs because they do not leverage the information within retrospective reports.

While AU estimates of IRs and IPs for ILI are not currently available, laboratory confirmed influenza is a nationally notifiable disease in AU.[35] In 2016 the age-specific rate estimates of laboratory confirmed influenza ranged from 24.67 to 123.72 per 10,000 population.[35] As expected, these estimates are lower than the ILI estimates from FluTracking because the reported number of cases represent only a proportion of the total cases in the community, that is, only those cases for which health care was sought, a test conducted and a diagnosis made, followed by a notification to health authorities and does not include the many other viral and bacterial causes of ILI. But the patterns in age-group specific estimates are similar, as are the weekly trends.

This study has several limitations. FluTracking has a highly engaged user population with a smaller proportion of missing reports compared with other systems. Because imputation methods may not be feasible with large amounts of missing data, other systems may need to select a cohort of highly engaged users before implementing MI methods. FluTracking is also unique because it provides users with the opportunity to complete missing surveys. This system accommodation not only adds approximately 10% more weekly reports, but also provides a way to estimate *δ* for imputation. Additionally, in our MNAR simulation model, because the value of *δ* is known, the resulting *δ*-MI model can be properly parameterized. In reality, this value is unknown, and the estimated value is based only on retrospective reports, which may be affected by recall bias. However, our sensitivity analysis shows that under modest changes in*δ*, for example, increasing *δ* (OR) from 0.3 (1.35) to 0.8 (2.22) or 1.3 (3.67), the *δ*-MI method still outperforms the method that assumes missing reports are non-ILI. Finally, only one value of *δ* was used for imputation for all age groups and sex. In the future, this parameter can be easily updated and adapted during an influenza season to provide real time, demographic-specific estimates. A Bayesian approach to estimation can also be used.

National estimates of influenza burden in the population are essential to understand the overall global burden of influenza disease.[37] Alternative data sources, such as FluTracking, have the potential to complement traditional sentinel systems by capturing a population not routinely included among the other healthcare-based systems and identifying healthcare seeking behaviors and testing rates among ill persons.[2] While these surveillance systems now play an important role in surveillance, data analysis needs to be handled carefully to draw appropriate and useful conclusions. The *δ*-MI method provides a straightforward, interpretable approach to appropriately handling missing data in participatory surveillance systems.

## Data Availability

Contact to request data. Data requests will be actioned within resource constraints.

## ABBREVIATIONS

AU: Australia
CI: Confidence Interval
COVID-19: Coronavirus 2019
FNY: Flu Near You
ILI: Influenza-Like Illness
IP: Incidence Proportion
IR: Incidence Rate
MAR: Missing at Random
MCAR: Missing Completely at Random
MI: Multiple Imputation
MICE: Multivariate Imputation by Chained Equations
MMWR: Morbidity and Mortality Weekly Report
MNAR: Missing Not at Random
NRMSE: Normalized Root Mean Square Error
US: United States of America
WHO: World Health Organization
WP: Weekly Prevalence

## ACKNOWLEDGEMENTS

The authors acknowledge all the participants who contributed their time and information to the FluTracking system.

## REFERENCES

1. Choi J, Cho Y, Shim E, Woo H. Web-based infectious disease surveillance systems and public health perspectives: a systematic review. BMC Public Health 2016; 16:1238. Available at: http://bmcpublichealth.biomedcentral.com/articles/10.1186/s12889-016-3893-0.

2. Smolinski MS, Crawley AW, Olsen JM, Jayaraman T, Crawley AW. Participatory Disease Surveillancel’.: Engaging Communities Directly in Reporting, Monitoring, and Responding to Health Threats Corresponding Authorl’.: 2017; 3.

3. Marquet RL, Bartelds AIM, van Noort SP, et al. Internet-based monitoring of influenza-like illness (ILI) in the general population of the Netherlands during the 2003-2004 influenza season. BMC Public Health 2006; 6:242. Available at: http://www.pubmedcentral.nih.gov/articlerender.fcgi?artid=1609118&tool=pmcentrez&rendertype=abstract.

4. Paolotti D, Carnahan A, Colizza V, et al. Web-based participatory surveillance of infectious diseases: The Influenzanet participatory surveillance experience. Clin. Microbiol. Infect. 2014; 20:17–21. Available at: http://dx.doi.org/10.1111/1469-0691.12477.

5. Moberley S, Carlson SJ, Durrheim DN, Dalton CB, DN. FluTracking: Weekly online community based surveillance of influenza-like illness in Australia, 2017 Annual Report. Communicable Diseases Intelligence 2019

6. Smolinski MS, Crawley AW, Baltrusaitis K, et al. Flu Near You: Crowdsourced Symptom Reporting Spanning 2 Influenza Seasons. Am. J. Public Health 2015; 105:2124–2130. Available at: http://ajph.aphapublications.org/doi/10.2105/AJPH.2015.302696.

7. Segal E, Zhang F, Lin X, et al. Building an international consortium for tracking coronavirus health status. Nat. Med. 2020; https://doi.org/10.1038/s41591-020-0929-x

8. Kullenberg C, Kasperowski D. What is citizen science? - A scientometric meta-analysis. PLoS One 2016; 11:1–16. Available at: http://dx.doi.org/10.1371/journal.pone.0147152.

9. Santillana M, Nguyen AT, Dredze M, Paul MJ, Nsoesie EO, Brownstein JS. Combining Search, Social Media, and Traditional Data Sources to Improve Influenza Surveillance. PLOS Comput. Biol. 2015; 11:e1004513. Available at: http://dx.plos.org/10.1371/journal.pcbi.1004513.

10. Perrotta D, Bella A, Rizzo C, Paolotti D. Participatory Online Surveillance as a Supplementary Tool to Sentinel Doctors for Influenza-Like Illness Surveillance in Italy. PLoS One 2017; 12:e0169801. Available at: http://dx.plos.org/10.1371/journal.pone.0169801.

11. van Noort SP, Codeço CT, Koppeschaar CE, van Ranst M, Paolotti D, Gomes MGM. Ten-year performance of Influenzanet: ILI time series, risks, vaccine effects, and care-seeking behaviour. Epidemics 2015; 13:28–36. Available at: http://linkinghub.elsevier.com/retrieve/pii/S1755436515000638.

12. Brownstein JS, Chu S, Marathe A, et al. Combining Participatory Influenza Surveillance with Modeling and Forecasting: Three Alternative Approaches. JMIR Public Heal. Surveill. 2017; 3:e83. Available at: http://publichealth.jmir.org/2017/4/e83/.

13. Moss R, Zarebski AE, Carlson SJ, McCaw JM. Accounting for Healthcare-Seeking Behaviours and Testing Practices in Real-Time Influenza Forecasts. Trop. Med. Infect. Dis. 2019; 4,12, doi:10.3390/tropicalmed4010012.

14. Patterson-Lomba O, Van Noort S, Cowling BJ, et al. Utilizing syndromic surveillance data for estimating levels of influenza circulation. Am. J. Epidemiol. 2014; 179:1394–1401.

15. Chunara R, Goldstein E, Patterson-lomba O, Brownstein JS. Estimating influenza attack rates in the United States using a participatory cohort. 2015;: 1–5.

16. Stockwell MS, Reed C, Vargas CY, et al. MoSAIC: Mobile surveillance for acute respiratory infections and influenza-like illness in the community. Am. J. Epidemiol. 2014; 180:1196– 1201.

17. Menni C, Valdes AM, Freidin MB, et al. Real-time tracking of self-reported symptoms to predict potential COVID-19. Nat. Med. 2020; https://doi.org/10.1038/s41591-020-0916-2

18. Carlson SJ, Durrheim DN, Dalton CB. FluTracking provides a measure of field influenza vaccine effectiveness, Australia, 2007-2009. Vaccine 2010; 28:6809–6810. Available at: http://dx.doi.org/10.1016/j.vaccine.2010.08.051.

19. Debin M, Colizza V, Blanchon T, Hanslik T, Turbelin C, Falchi A. Effectiveness of 2012 – 2013 influenza vaccine against influenza-like illness in general population Estimation in a French web-based cohort. 2014; 10:536–543.

20. Tilston NL, Paolotti D, Ealden T. Internet-based surveillance of Influenza-like-illness in the UK during the 2009 H1N1 influenza pandemic. BMC Public Health 2010; 10:1–9.

21. Peppa M, John Edmunds W, Funk S. Disease severity determines health-seeking behaviour amongst individuals with influenza-like illness in an internet-based cohort. BMC Infect. Dis. 2017; 17:1–13.

22. Baltrusaitis K, Reed C, Sewalk K, Brownstein JS, Crawley AW, Biggerstaff M. Health-care seeking behavior for respiratory illness among Flu Near You participants in the United States during the 2015-16 through 2018-19 influenza season. JID. 2020;

23. Chunara R, Wisk LE, Weitzman ER. Denominator Issues for Personally Generated Data in Population Health Monitoring. Am. J. Prev. Med. 2017; 52:549–553. Available at: http://dx.doi.org/10.1016/j.amepre.2016.10.038.

24. Baltrusaitis K, Santillana M, Crawley AW, Chunara R, Smolinski M, Brownstein JS. Determinants of Participants’ Follow-Up and Characterization of Representativeness in Flu Near You, A Participatory Disease Surveillance System. JMIR public Heal. Surveill. 2017; 3:e18. Available at: http://publichealth.jmir.org/2017/2/e18/%0Ahttp://www.ncbi.nlm.nih.gov/pubmed/28389417.

25. FluTracking. Available at: https://info.FluTracking.net/. Accessed 19 April 2021.

26. (WHO) WHO. A Manual for Estimating Disease Burden Associated with Seasonal Influenza. http://apps.who.int/iris/bitstream/10665/178801/1/9789241549301_eng.pdf?ua=1&ua=1. Accessed November 28, 2017. Who 2015;: 124. Available at: http://www.who.int/influenza/resources/publications/manual_burden_of_disease/en/.

27. Kirkwood BR, Sterne J. Essentials of Medical Statistics. 2nd ed. Blackwell Science Ltd, 2003.

28. Giesecke J. Modern Infectious Disease Epidemiology. 2nd ed. London, United Kingdom: Taylor & Francis Ltd, 2002.

29. Rubin DB. Multiple Imputation for Nonresponse in Surveys. New York: John Wiley and Sons, 2004.

30. Leacy FP, Floyd S, Yates TA, White IR. Analyses of sensitivity to the missing-at-random assumption using multiple imputation with delta adjustment: Application to a tuberculosis/HIV prevalence survey with incomplete HIV-status data. Am. J. Epidemiol. 2017; 185:304–315.

31. R Core Team (R Foundation for Statistical Computing). R: A Language and Environment for Statistical Computing. 2016; Available at: https://www.r-project.org/.

32. Buuren S van, Groothuis-Oudshoorn K. miceli⍰: Multivariate Imputation by Chained Equations in R. J. Stat. Softw. 2011; 45. Available at: http://www.jstatsoft.org/v45/i03/.

33. AUSTRALIAN INFLUENZA Laboratory Confirmed Influenza Activity. 2016;: 1–10. Available at: http://www.health.gov.au.

34. Lui D, Mitchell L, Cope RC, Carlson SJ, and Ross JV. Elucidating User Behaviors in a Digital Health Surveillance System to Correct Prevalence Estimates. Epidemics. 2020; 33

35. Sullivan SG, Kate Pennington JR, Franklin LJ, et al. A Summary of Influenza Surveillance Systems in Australia, 2015. Commun. Dis. Intell. 2016;: 1–51. Available at: http://www.health.gov.au/internet/main/publishing.nsf/Content/cda-surveil-ozflu-flucurr.htm/$File/Influenza-Surveillance-Systems-Paper.pdf.

36. National Notifiable Diseases Surveillance System. 2016. Available at: http://www.health.gov.au/nndssdata. Accessed 15 July 2018.

37. Yang W, Lipsitch M, Shaman J. Inference of seasonal and pandemic influenza transmission dynamics. Proc. Natl. Acad. Sci. 2015; 112:2723–2728. Available at: http://www.pnas.org/content/112/9/2723.abstract

