## Supplemental for "Missing data matters in participatory syndromic surveillance systems: comparative evaluation of missing data methods when estimating disease burden": Flutracking_missing_supplementary_30APR2021.docx

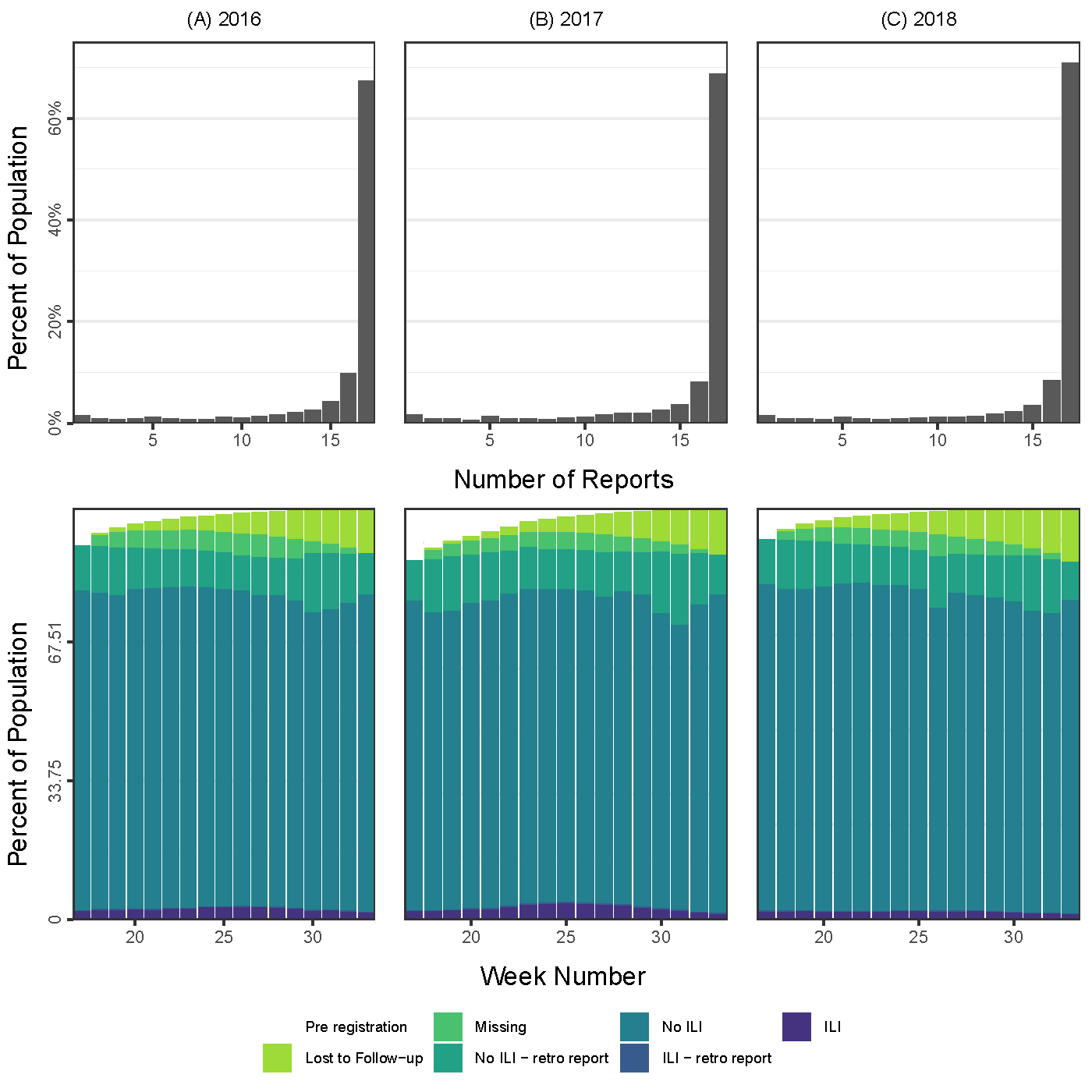

**Supplemental Figure 1.** Distributions of the number of FluTracking participant reports and histograms of FluTracking participant reporting habits during the (A) 2016, (B) 2017, and (C) 2018 reporting periods.

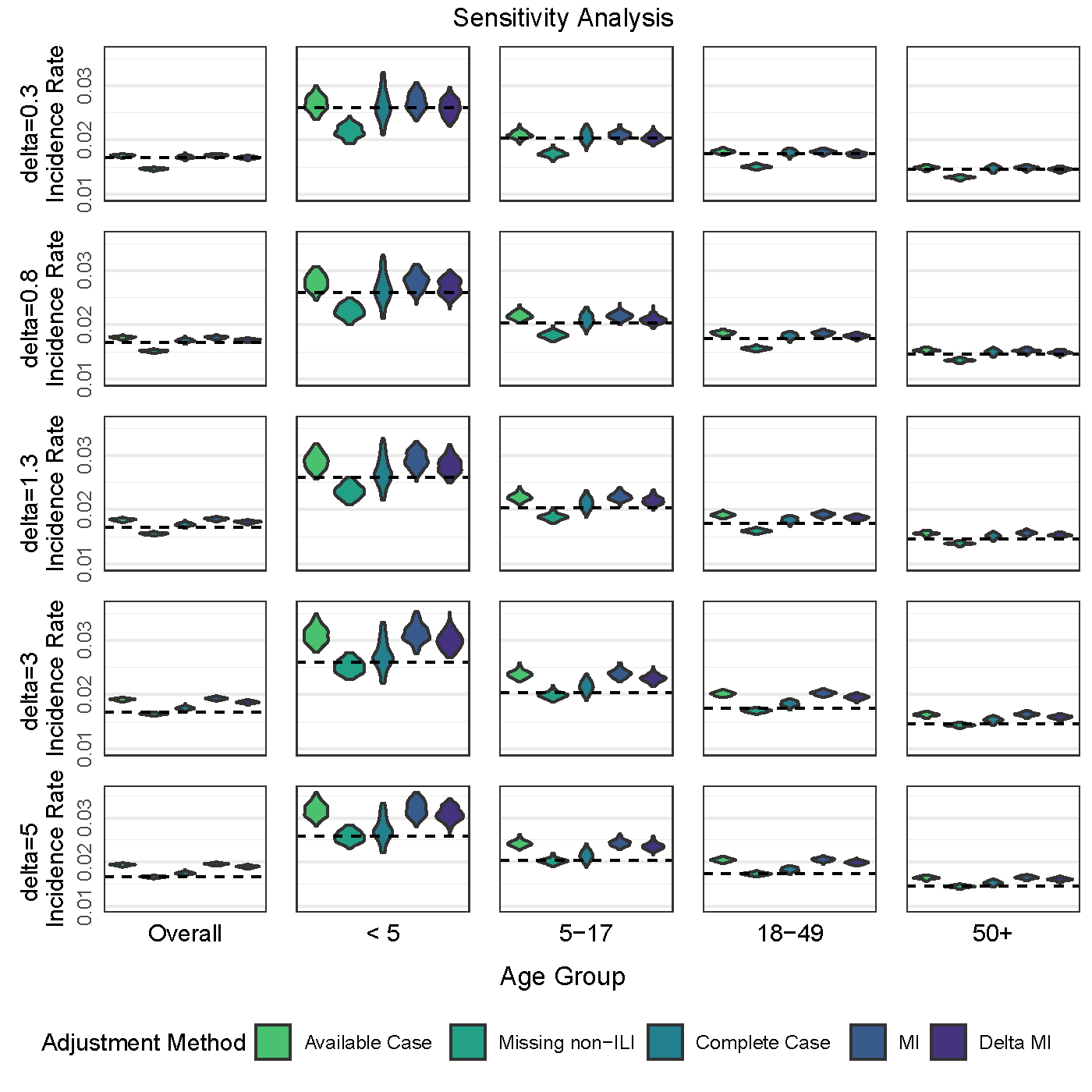

**Supplemental Figure 2.** Distributions of overall and age group specific Incidence Ratios simulated under five Missing Not at Random (MNAR) scenarios. Dotted line represents the original simulated parameter.

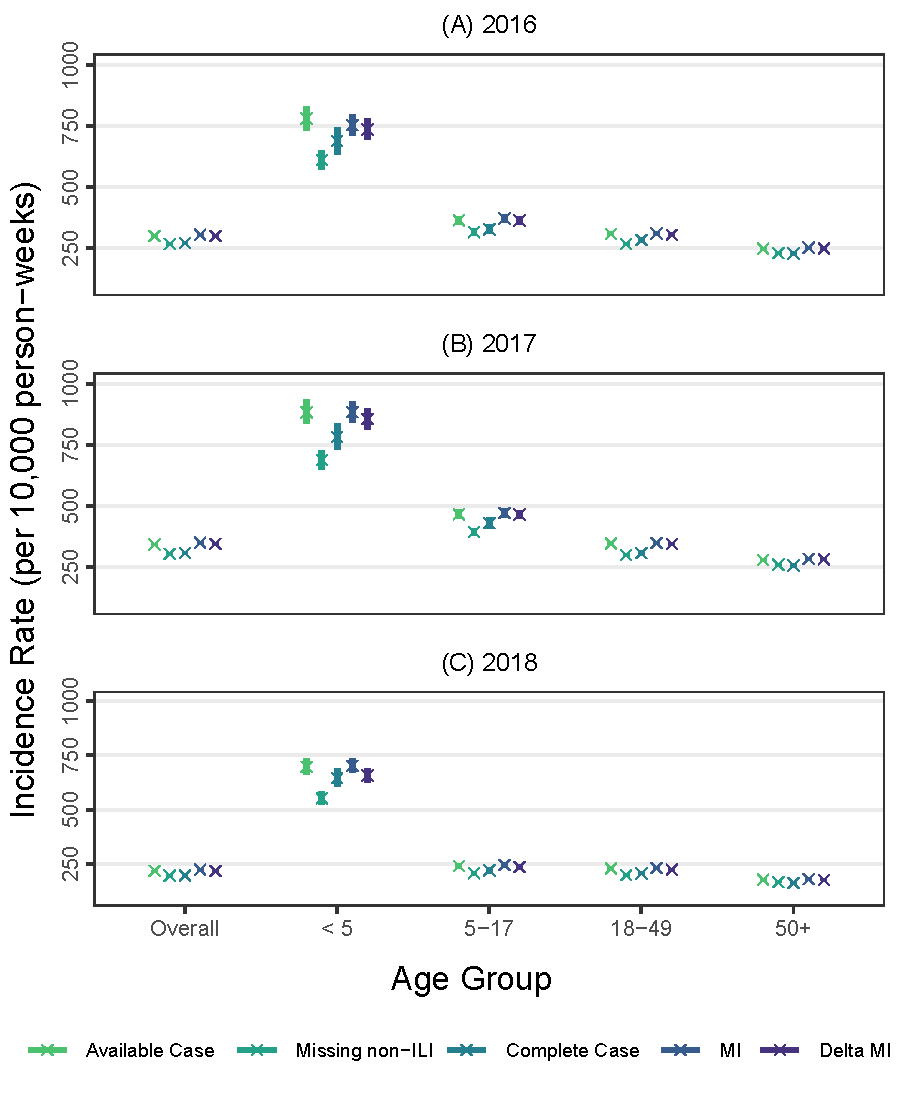

**Supplemental Figure 3.** Overall and age group specific Incidence Rates and 95% Confidence Intervals for FluTracking, expressed as number of ILI reports per 10,000 person weeks, for the (A) 2016, (B) 2017, and (C) 2018 influenza seasons.

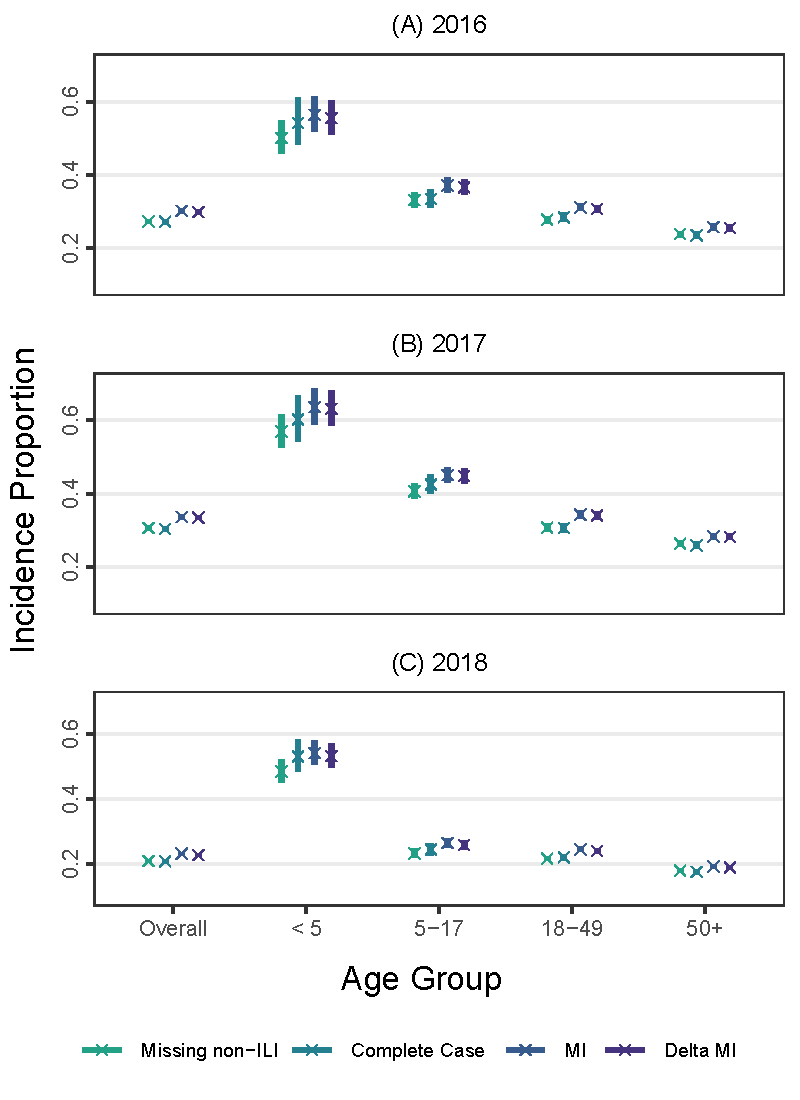

**Supplemental Figure 4.** Overall and age group specific Incidence Proportions and 95% Confidence Intervals for FluTracking, expressed as percent of population reporting ILI at least once, for the (A) 2016, (B) 2017, and (C) 2018 influenza seasons.

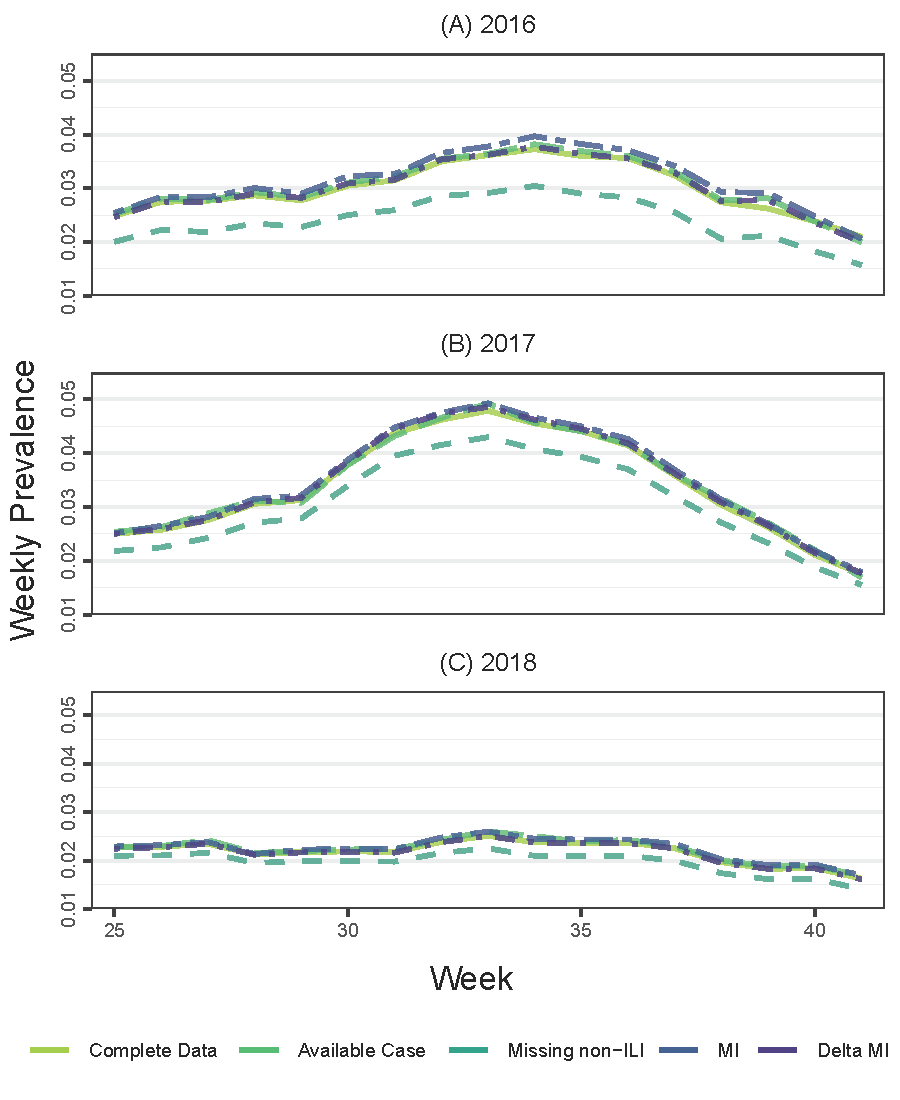

**Supplemental Figure 5.** Weekly Prevalence for FluTracking for the (A) 2016, (B) 2017, and (C) 2018 influenza seasons.

**Supplemental Table 1.** Overall and age group specific Incidence Rates and 95% Confidence Intervals for FluTracking, expressed as number of ILI reports per 10,000 person weeks, by influenza season.

| **Season** | **Age Group** | **Missing Data Method** | | | | |
| --- | --- | --- | --- | --- | --- | --- |
|  |  | **Available Case** | **Assume missing are non-ILI** | **Complete Case** | **Imputation** | $\boldsymbol{\delta}$ **Imputation** |
| 2016 | Overall | 299.4  (294.4, 304.5) | 267.1  (262.6, 271.7) | 271.9  (266.3, 277.6) | 305.3  (300.6, 310.2) | 299.6  (294.9, 304.4) |
|  | <5 | 780.4  (733.5, 830.3) | 610.2  (573.5, 649.3) | 688.4  (635.4, 745.8) | 753.4  (712.5, 796.6) | 736.1  (695.6, 778.8) |
|  | 5-17 | 364.2  (347.8, 381.3) | 315.2  (301.0, 330.1) | 327.2  (308.5, 346.9) | 370.6  (355.2, 386.7) | 362.8  (347.6, 378.7) |
|  | 18-49 | 308.4  (300.3, 316.8) | 267.7  (260.7, 275.0) | 283.5  (274.3, 293.1) | 311.0  (303.4, 318.8) | 304.6  (297.1, 312.4) |
|  | 50+ | 248.0  (241.4, 254.7) | 230.0  (223.9, 236.3) | 228.7  (221.4, 236.2) | 252.1  (245.7, 258.7) | 248.3  (242.0, 254.8) |
| 2017 | Overall | 340.8  (335.7, 346.0) | 303.1  (298.6, 307.7) | 307.1  (301.5, 312.8) | 348.7  (343.8, 353.6) | 344.3  (339.5, 349.3) |
|  | <5 | 882.6  (834.8, 933.1) | 687.4  (650.2, 726.7) | 782.1  (730.0, 837.8) | 883.6  (841.3, 928.0) | 855.5  (813.9, 899.3) |
|  | 5-17 | 464.5  (446.8, 482.9) | 392.3  (377.4, 407.9) | 429.1  (409.1, 450.1) | 470.5  (454.1, 487.5) | 464.2  (448.0, 481.1) |
|  | 18-49 | 345.7  (337.2, 354.4) | 298.6  (291.2, 306.1) | 306.1  (296.8, 315.6) | 347.9  (339.9, 355.9) | 343.6  (335.7, 351.6) |
|  | 50+ | 278.7  (272.2, 285.4) | 258.8  (252.7, 265.0) | 255.3  (248.1, 262.7) | 283.2  (276.8, 289.6) | 280.8  (274.5, 287.3) |
| 2018 | Overall | 218.5  (214.9, 222.0) | 196.2  (193.0, 199.4) | 197.6  (193.8, 201.6) | 224.1  (220.7, 227.5) | 217.5  (214.1, 220.8) |
|  | <5 | 696.7  (661.3, 734.0) | 552.3  (524.3, 581.9) | 644.8  (604.5, 687.7) | 701.9  (670.2, 735.2) | 657.8  (627.2, 690.0) |
|  | 5-17 | 240.6  (229.6, 252.2) | 207.3  (197.8, 217.2) | 220.6  (208.2, 233.6) | 244.5  (234.2, 255.3) | 236.1  (225.9, 246.7) |
|  | 18-49 | 229.6  (223.5, 235.9) | 198.4  (193.1, 203.8) | 206.7  (200.0, 213.7) | 231.8  (226.1, 237.6) | 224.1  (218.5, 229.9) |
|  | 50+ | 177.5  (173.1, 182.1) | 166.7  (162.6, 171.0) | 163.0  (158.2, 167.9) | 179.8  (175.5, 184.2) | 177.0  (172.7, 181.4) |

**Supplemental Table 2.** Overall and age group specific Incidence Proportions and 95% Confidence Intervals for FluTracking, expressed as percent of population reporting ILI at least once, by influenza season.

| **Season** | **Age Group** | **Missing Data Method** | | | |
| --- | --- | --- | --- | --- | --- |
|  |  | **Assume missing are non-ILI** | **Complete Case** | **Imputation** | $\boldsymbol{\delta}$**-Imputation** |
| 2016 | Overall | 27.33  (26.74, 27.93) | 27.22  (26.49, 27.97) | 30.20  (29.58, 30.83) | 29.82  (29.21, 30.45) |
|  | <5 | 50.05  (45.78, 54.73) | 54.21  (48.19, 60.98) | 56.37  (51.82, 61.31) | 55.50  (50.99, 60.41) |
|  | 5-17 | 33.10  (31.22, 35.09) | 33.45  (31.01, 36.08) | 37.12  (35.13, 39.23) | 36.60  (34.62, 38.69) |
|  | 18-49 | 27.79  (26.86, 28.76) | 28.40  (27.19, 29.66) | 31.16  (30.17, 32.18) | 30.69  (29.71, 31.71) |
|  | 50+ | 23.85  (23.04, 24.69) | 23.53  (22.57, 24.53) | 25.77  (24.92, 26.64) | 25.53  (24.69, 26.40) |
| 2017 | Overall | 30.63  (30.04, 31.24) | 30.31  (29.58, 31.05) | 33.60  (32.98, 34.24) | 33.42  (32.80, 34.06) |
|  | <5 | 56.97  (52.60, 61.69) | 60.16  (54.31, 66.65) | 63.56  (58.94, 68.54) | 63.08  (58.48, 68.04) |
|  | 5-17 | 40.62  (38.65, 42.70) | 42.48  (39.90, 45.22) | 45.00  (42.92, 47.18) | 44.77  (42.69, 46.94) |
|  | 18-49 | 30.72  (29.75, 31.71) | 30.63  (29.43, 31.89) | 34.22  (33.20, 35.27) | 34.02  (33.00, 35.06) |
|  | 50+ | 26.33  (25.53, 27.14) | 25.82  (24.88, 26.79) | 28.29  (27.47, 29.14) | 28.16  (27.34, 29.01) |
| 2018 | Overall | 20.95  (20.53, 21.39) | 20.81  (20.29, 21.33) | 23.13  (22.68, 23.58) | 22.71  (22.27, 23.17) |
|  | <5 | 48.34  (44.95, 51.98) | 53.02  (48.33, 58.18) | 54.00  (50.41, 57.84) | 53.08  (49.53, 56.89) |
|  | 5-17 | 23.22  (21.92, 24.6) | 24.39  (22.71, 26.20) | 26.40  (25.01, 27.87) | 25.78  (24.40, 27.23) |
|  | 18-49 | 21.67  (20.95, 22.41) | 22.05  (21.15, 23.00) | 24.46  (23.70, 25.25) | 23.96  (23.20, 24.73) |
|  | 50+ | 17.95  (17.39, 18.53) | 17.55  (16.90, 18.22) | 19.19  (18.61, 19.79) | 18.93  (18.35, 19.52) |
